# Early identification of advanced chronicity (MACA) patients using Machine Learning models: a population-based predictive approach for proactive care stratification

**DOI:** 10.64898/2026.06.16.26355503

**Authors:** Miguel Boubeta, Marc Moreno Ariño, Oscar Duems Noriega, Marina Roig Soronellas, Julia Veríssimo Guillén, Ingrid Bullich Marín, Carina Sanz Blazquez, Joana Barrio Medina, Montserrat López Postigo, Luis Lorenzo, Marcos Montaña-Méndez, Cristóbal Bernardo-Castiñeira, María Dolores López Lores, Víctor Borrás-Marco, Sandra Posas Paradera

## Abstract

Early identification of patients with advanced chronic conditions (MACA) remains a critical challenge in clinical practice, often relying on retrospective criteria or clinical judgment, which may delay timely and personalized intervention. The increasing availability of electronic health records (EHR) enables the application of Machine Learning (ML) techniques to support more proactive detection. This study aimed to develop and internally validate a ML-based approach for the identification of MACA patients using collected data from Hospital Universitario Parc Taulí (Sabadell, Spain). A retrospective observational study was conducted using a sample of 163 patients. A total of 80 candidate variables were extracted, including clinical, functional, and healthcare utilization indicators. Feature selection methods were applied, reducing the dataset to ten key predictors. Fourteen supervised classification algorithms were evaluated, including linear, probabilistic, and ensemble methods. Model performance was evaluated using various metrics like accuracy, precision, recall, F1-score, and area under the receiver operating characteristic curve (AUC-ROC). The final cohorts include 80 MACA patients and 83 controls. The bagging classifier achieved the most consistent performance with a sensitivity of 0.91 and an AUC of 0.90. Key predictors include absolute dependency, advanced frailty, functional decline, and healthcare utilization indicators. Cross-validation (CV) confirmed the stability of model performance, with mean AUC values exceeding 0.95. These findings highlight the potential of ML-based tools for early detection and the high discriminative capacity for identifying MACA patients.

## 1. Introduction

The global rise in chronic diseases represents a structural challenge for modern healthcare systems, driven by population aging, improved survival rates, and the increasing prevalence of multimorbidity. Patients with multiple chronic diseases account for a disproportionate share of healthcare utilization, costs, and mortality, particularly in high-income countries (Barnet et al., 2012; Violan et al., 2014). Within this heterogeneous population, a subgroup of patients with advanced chronic conditions, exhibits heightened clinical vulnerability, characterized by functional decline, high symptom burden, frequent hospital admissions, and limited life expectancy (Jiménez et al., 2021; Gómez-Batiste et al., 2013; Lynn & Adamson, 2003; Murray et al., 2005). Accurate and early identification of these patients is essential to enable timely implementation of palliative care approaches, advance care planning, and coordinated patient-centred interventions (Temel et al., 2010; Smith et al., 2016). However, current identification strategies still have room for improvement. Tools such as NECPAL (Gómez-Batiste et al., 2013) and comorbidity-based indices like the Charlson Comorbidity Index (Charlson et al., 1987) are widely used but present important limitations: reliance on clinical judgement, limited sensitivity for early-stage identification and insufficient capacity to capture the multidimensional and dynamic nature of advanced chronicity (Bernabeu-Wittel et al., 2011; Boyd et al., 2005). Additionally, prognostic uncertainty in non-oncological chronic diseases further complicates timely recognition of patients approaching advanced stages (Murray et al., 2005; Lunney et al., 2003).

In parallel, the increasing availability of large-scale EHRs has catalysed the application of ML methods for clinical prediction. Compared to traditional statistical approaches, ML models offer enhanced capacity to model complex relationships and interactions across high-dimensional datasets (Obermeyer & Emanuel, 2016; Rajkomar et al., 2018). Classification algorithms such as random forest, gradient boosting machines, and penalized regression techniques have demonstrated superior predictive performance in diverse healthcare applications, including mortality prediction, hospital readmissions, and disease progression (Kansagara et al., 2011; Futoma et al., 2015). Several studies have explored ML-based risk stratification in chronic disease populations. Predictive models leveraging administrative and clinical data have shown improved discrimination for identifying high-risk patients compared to rule-based systems (Billings et al., 2012; Hippisley-Cox & Coupland, 2017). Similarly, Deep Learning (DL) approaches applied to EHR data have demonstrated high accuracy predicting clinical outcomes across large patient cohorts (Rajkomar et al., 2018). Nevertheless, most of these studies focus on outcomes such as short-term mortality, emergency admissions, or specific complications of the disease, rather than addressing the broader concept of advanced chronicity (Krumholz, 2014; Goldstein et al., 2017).

The identification of MACA patients requires integration of multiple dimensions, including multimorbidity, functional status, healthcare trajectories, and markers of clinical deterioration. Traditional models often fail to capture this complexity, whereas ML approaches offer a promising alternative by enabling data-driven phenotyping and pattern recognition (Miotto et al., 2016; Shickel et al., 2018). However, significant challenges remain, including model interpretability, risk of bias, lack of external validation, and limited integration into clinical workflows (Wynants et al., 2020; Kelly et al., 2019). Moreover, concerns regarding algorithmic fairness and generalizability highlight the need for rigorous methodological standards in predictive modelling studies (Obermeyer et al., 2019). Despite the growing body of literature on Artificial Intelligence (AI) and ML in healthcare, there is still a notable gap in studies specifically targeting the early detection of advanced chronicity in real-world population-based settings. Existing approaches to identifying MACA patients are often retrospective or triggered by late-stage clinical events, limiting their utility for proactive care planning (Gómez-Batiste et al., 2013).

The present study aims to address these gaps by developing and validating ML classification models for the early identification of patients with advanced chronic conditions within a population-based cohort. We hypothesize that ML models can outperform conventional identification strategies by leveraging collected healthcare data to detect complex, latent patterns associated with disease progression and clinical deterioration. In addition, we aim to assess model calibration, interpretability, and potential clinical utility in supporting proactive care stratification. By integrating advanced analytical techniques with clinically meaningful endpoints, this study seeks to contribute to the development of scalable, robust, and clinically applicable tools for the early identification of MACA patients, ultimately enabling more efficient, proactive and patient-centred care.

The remainder of this article is structured as follows. The Materials and Methods section describes the study design, data sources, population selection criteria, outcome definition, feature engineering process, and the development and validation of the ML models, including performance metrics and evaluation strategy. Model performance, robustness and sensitivity analysis are covered in Results section. The Discussion section critically interprets these findings, addressing methodological strengths, potential sources of bias, limitations related to data and generalizability, and implications for clinical practice. Finally, the Conclusions section summarizes the main contributions of the study and outlines directions for future research.

## 2. Materials and Methods

This study was designed as a retrospective observational cohort analysis conducted at Hospital Universitario Parc Taulí (Sabadell, Spain). The study population comprised patients admitted to the hospital who were considered candidates for assessment of advanced chronicity. To preserve clinical coherence and avoid structural heterogeneity, patients admitted to Paediatrics, Obstetrics and Gynaecology, Mental Health, and Intensive Care Units were excluded, as these patients are characterised by different care trajectories and are not aligned with the operational definition of advanced chronicity addressed in this study. Although it is known that the prevalence of advanced chronic disease increases with age, no age restrictions have been applied to capture the broadest clinical spectrum of the disease. However, patients managed within the Paediatrics service were excluded from the study population. The final dataset includes a representative sample of both populations, MACA and non-MACA patients. The study sample comprised 163 patients, of whom 80 (49.1%) were classified as MACA and 83 (50.9%) as non-MACA, with all patient data pseudo-anonymised prior to analysis in accordance with data protection regulations to ensure confidentiality.

All data were extracted from the Hospital Parc Taulí EHRs, including structured clinical variables, diagnostic information, functional assessments, and healthcare utilization indicators. Among all hospital admissions, approximately 15% correspond to chronic patients, within whom 3.5% exhibit complex chronicity and 1.5% are classified as MACA. The response variable was defined as MACA status (binary, 1 if the patient is MACA and 0 otherwise), established according to the criteria of clinical experts and confirmed with the NECPAL 4.0 tool. Initially, experts identified a set of 173 variables, organised into 24 conditions, each grouping variables with shared characteristics. The list of conditions includes: (1) identified palliative care needs, (2) advanced frailty, (3) intermediate frailty, (4) advanced dementia, (5) moderate dementia, (6) extreme age, (7) very high risk according to prognostic scales, (8) absolute dependence, (9) severe dependence or functional decline, (10) moderate dependence, (11) complex chronicity profile, (12) severe or progressing cancer, (13) severe or progressing lung disease, (14) severe or progressing heart disease, (15) severe or progressing liver disease, (16) severe or progressing kidney disease, (17) severe or progressing cerebrovascular disease, (18) severe or progressing neurodegenerative disease, (19) use of specific resources, (20) high use of resources, (21) complexity in prescribing, (22) hard geriatric syndromes, (23) medium geriatric syndromes, and (24) soft geriatric syndromes.

Due to the complexity and high resource demand for extracting data from the hospital’s EHRs, finally, of the 173 variables initially identified, 80 (62 clinical variables and 18 conditions) were selected as explanatory features for training the ML models. To enhance clinical plausibility and reduce the risk of spurious associations, domain-informed hierarchical dependencies between some clinical conditions were explicitly incorporated into the feature space. These include predefined progressions between related conditions (condition 2 to 3, condition 4 to condition 5, and condition 8 through condition 9 to condition 10). During model development, variables within condition 1, reflecting prior engagement with specialized palliative care resources, were found to disproportionately drive model predictions. Consequently, these variables were excluded, prioritizing predictors related to underlying clinical complexity, functional decline, and healthcare utilization. This strategy aimed to enhance the model’s ability to identify previously unrecognized MACA patients and improve its practical value as a screening tool.

Data processing followed a structured pipeline aimed at ensuring data quality and model robustness. Variables and observations with excessive missingness were excluded, and the remaining missing values were handled using simple imputation strategies (numerical variables were imputed using the mean and categorical variables assigned a value of 0 reflecting absence of the condition). Subsequently, categorical variables were transformed using the one hot encoding strategy and continuous variables were standardised to zero mean and unit variance. To address the high dimensionality, three feature selection strategies (k-best, low variance, and recursive feature elimination) were applied to prioritise variables with strongest statistical association with the response variable while preserving clinical interpretability.

In total, 13 supervised classification algorithms were evaluated, encompassing both conventional statistical approaches and more advanced ensemble methods. The first group comprises linear models, including logistic regression, linear discriminant analysis, naïve Bayes and support vector machines, whereas the second group includes non-linear methods, such as decision trees, gradient boosting (e.g., xgboost, catboost and light gradient boosting machine), and bagging-based methods (e.g., bagging classifier, random forest and extra trees). Hyperparameter tuning was performed ensuring robust optimization while minimizing overfitting. Model selection was driven not only by predictive performance but also by considerations of clinical relevance and applicability.

Model performance was assessed using a dual validation strategy. An initial hold-out approach was used, with a 75/25 split between training and testing sets respectively to estimate generalization performance. This was complemented by a stratified 5-fold CV to enhance robustness and mitigate variance associated with the relatively small sample size. Performance metrics include accuracy, precision, recall (sensitivity), F1-score and AUC-ROC. Given the primary objective of early identification, model evaluation has prioritized sensitivity, as false negatives may delay appropriate care planning and result in adverse clinical consequences.

## 3. Results

Model performance was evaluated on an independent set of 41 patients. Table 1 presents the evaluation results for each model using the variables provided by the feature selection method. Among the 80 auxiliary variables, the feature selection method has identified 10: COND8 (absolute dependency), COND9 (severe dependence or functional decline), COND10 (moderate dependence), COND19 (use of specific resources), COND22 (hard geriatric syndromes), COND23 (medium geriatric syndromes), ccp (complex chronic patient identifier), coat_palliative (comprehensive outpatient assessment team palliative care), hcsp (admission or intervention by teams from the home care and support program) and neuroleptics (prescription of neuroleptics).

**Table 1:** Summary of performance metrics for the 13 models evaluated on the test set (n = 42), including confusion matrix components (TN, TP, FN, FP), accuracy (acc.), precision (prec.), recall (sensitivity), specificity, F1-score, and area under the ROC curve (AUC).

| Model | TN | TP | FN | FP | Acc. | Prec. | Recall | Specificity | F1-score | AUC |
| --- | --- | --- | --- | --- | --- | --- | --- | --- | --- | --- |
| Logistic regression | 20 | 15 | 6 | 1 | 0.833 | 0.938 | 0.714 | 0.952 | 0.811 | 0.876 |
| Catboost | 20 | 10 | 11 | 1 | 0.714 | 0.909 | 0.476 | 0.952 | 0.625 | 0.814 |
| Bagging | 19 | 19 | 2 | 2 | 0.905 | 0.905 | 0.905 | 0.905 | 0.905 | 0.902 |
| Gradient boosting | 20 | 12 | 9 | 1 | 0.762 | 0.923 | 0.571 | 0.952 | 0.706 | 0.796 |
| Linear discriminant analysis | 21 | 13 | 8 | 0 | 0.810 | 1.000 | 0.619 | 1.000 | 0.765 | 0.875 |
| AdaBoost | 20 | 14 | 7 | 1 | 0.810 | 0.933 | 0.667 | 0.952 | 0.778 | 0.798 |
| Random forest | 18 | 16 | 5 | 3 | 0.810 | 0.842 | 0.762 | 0.857 | 0.800 | 0.882 |
| XGBoost | 19 | 17 | 4 | 2 | 0.857 | 0.895 | 0.810 | 0.905 | 0.850 | 0.889 |
| LGBM | 19 | 17 | 4 | 2 | 0.857 | 0.895 | 0.810 | 0.904 | 0.850 | 0.912 |
| Linear SVC | 20 | 18 | 3 | 1 | 0.905 | 0.947 | 0.857 | 0.952 | 0.900 | 0.925 |
| Decision tree | 19 | 19 | 2 | 2 | 0.905 | 0.905 | 0.905 | 0.905 | 0.905 | 0.912 |
| Extra trees | 19 | 18 | 3 | 2 | 0.881 | 0.900 | 0.857 | 0.904 | 0.878 | 0.881 |
| Naive Bayes | 20 | 7 | 14 | 1 | 0.643 | 0.875 | 0.333 | 0.952 | 0.483 | 0.775 |

As can be seen in Table 1, among the 13 supervised classification algorithms assessed, bagging and decision tree classifiers achieved the highest overall performance. Both models exhibit very similar behaviour in the test set and achieve solid and identical values in the analysed performance metrics (confusion matrix, accuracy, precision, recall. F1-score and AUC), demonstrating that the approaches are competitive.

Classification results for both models showed 19 true positives and true negatives, and 2 false positives and false negatives. Both models yielded an accuracy, precision, recall (sensitivity) and F1-score of 90.5%. As can be seen in Figure 1, the AUC-ROC was 90.2% (bagging) and 91.2% (decision tree). Other alternative models also demonstrated high discriminatory capacity as linear SVC (F1-score 0.9; AUC 0.925), LGBM (F1-score 0.85; AUC 0.912) or extra trees (F1-score 0.878; AUC 0.881), although they have more FN and none surpassed the performance (in terms of F1-score) of bagging and extra trees classifiers.

**Figure 1:**
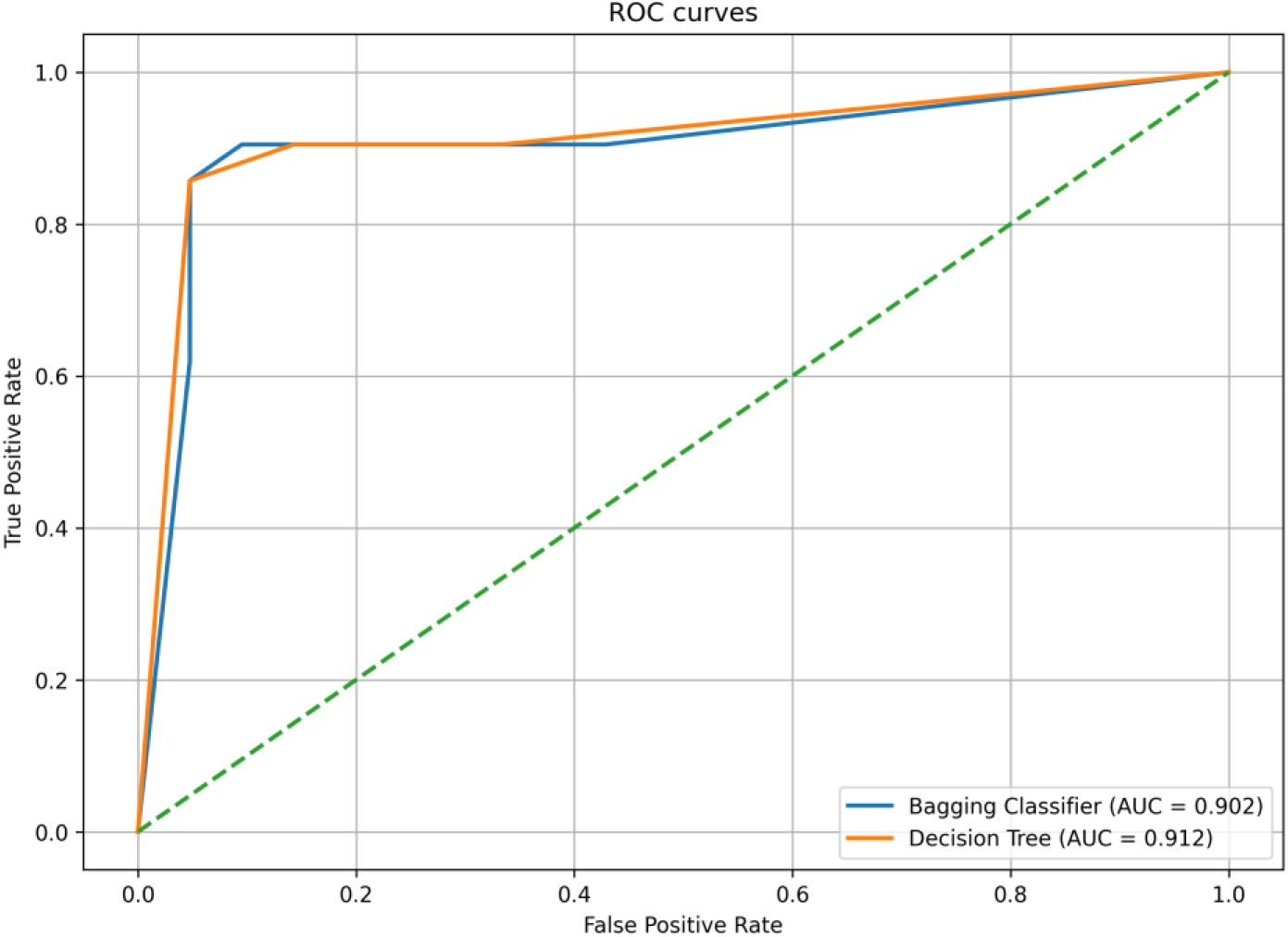
ROC curve for bagging and decision tree models.

These results indicate that, considering the metrics obtained on the test set, there are no significant differences between the bagging and decision tree models, both being able to effectively identify MACA patients.

To assess robustness, a stratified 5-fold CV procedure was applied to bagging and extra trees classifiers. Table 2 presents the results evaluated for each k-fold. Cross-validation results show a consistent behaviour for both models across the different folds, with limited variations and within expected ranges. However, the bagging performance metrics show slightly better results. The means of F1-score and AUCs across the 5-folds were 0.919 and 0.953 for bagging and 0.891 and 0.893 for decision tree, respectively.

**Table 2:**
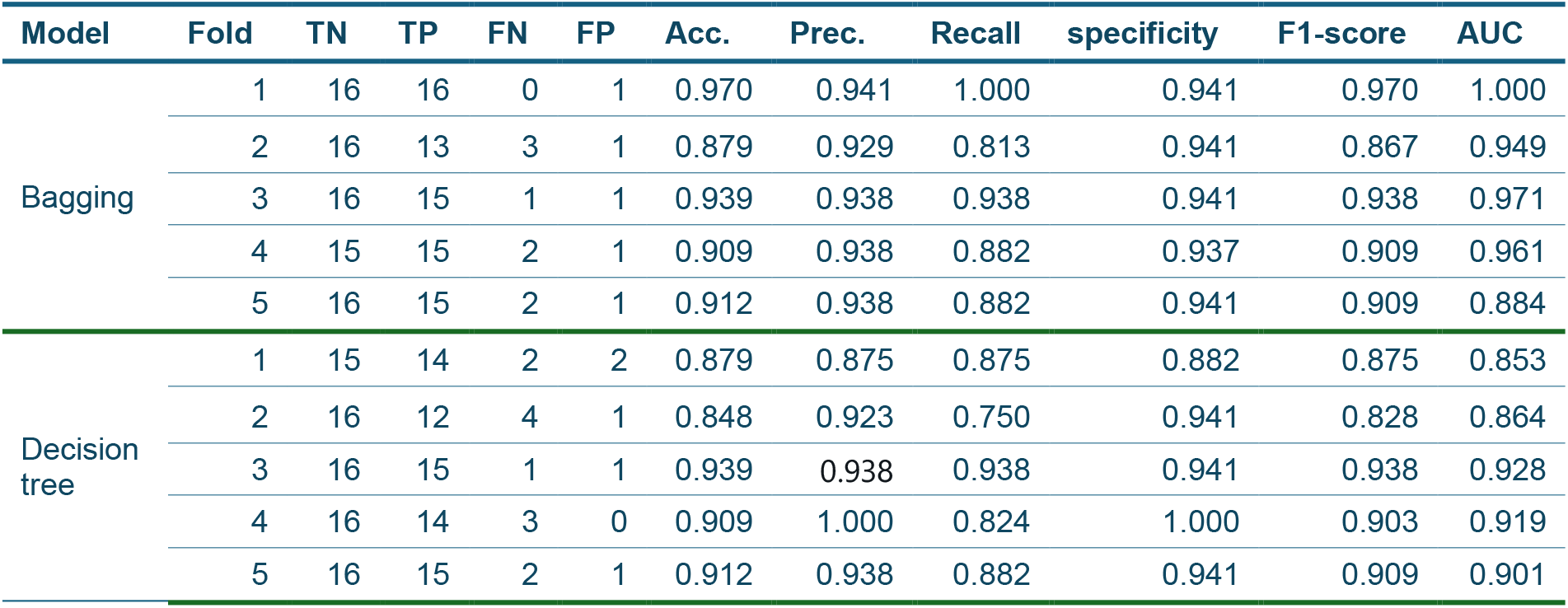
Summary of performance metrics for bagging and decision tree models evaluated on a 5-fold CV procedure, including confusion matrix components (TN, TP, FN, FP), accuracy (acc.), precision (prec), recall (sensitivity), specificity, F1-score, and area under the ROC curve (AUC).

This behaviour demonstrates that bagging classifier exhibits more robust behaviour, since it does not depend on a specific share and has adequate generalizability, reinforcing its suitability for the early identification of MACA patients. Feature importance analysis based on the bagging classifier identified absolute dependency (COND8) and use of specific resources (COND19) as the most influential predictors with relative importance weights of 0.318 and 0.262, respectively. The remaining selected variables contributed with lower but consistent weights.

## 4. Discussion

This study demonstrates that ML-based classification models can achieve high discriminative performance for the identification of patients with advanced chronic conditions in a real-world hospital setting. A parsimonious set of clinically interpretable variables reflecting functional decline, dependency and healthcare utilization was sufficient to yield a competitive discrimination in the test set. These findings reinforce de hypothesis that advanced chronicity is related to the convergence of functional decline, the complexity of care needs, and de cumulative burden of morbidity.

From a methodological standpoint, bagging and decision tree classifiers achieves a high discriminative ability compared to both linear and other ensemble approaches. While decision tree, LGBM, linear SVC and extra trees also demonstrated a competitive performance, the bagging classifier consistently achieved the most favourable balance between sensitivity and robustness. This finding suggests that aggregating multiple weak learners may better capture complex non-linear interactions compared to simpler predictors. At the same time, the observed performance must be interpreted cautiously given the limited sample size, which may still constrain generalizability and increase the risk of overfitting despite internal validation procedures. Importantly, although ensemble methods such as bagging may offer incremental improvements in predictive performance, they introduce additional challenges in terms of interpretability and clinical transparency.

The variables strongly associated with MACA status (absolute dependency, use of specific resources, advanced frailty and functional decline) are consistent with established conceptual models of advanced chronic disease trajectories. Prior work (Murray et al., 2005; Boyd et al., 2005) has shown that functional status and loss of autonomy are among the most robust predictors of mortality and healthcare utilization in multimorbid populations. Similarly, markers of healthcare utilization, such as enrolment in home-based care programs or indirect indicators of resource intensity, likely reflect latent dimensions of clinical complexity not reflected in diagnostic coding. In this context, the integration of clinical logic through hierarchical dependencies between conditions represents a methodological strength, as it aligns the modelling process with real-world disease progression patterns rather than purely statistical associations. Furthermore, the integration of relevant information extracted from unstructured data sources (e.g., nurse notes) through natural language processing has made it possible to achieve a high level of sensitivity and to adequately reflect clinical reasoning.

These results align with existing evidence on the use of ML for risk stratification in complex patient populations. Prior studies (Rajkomar et al., 2018) have reported improved predictive performance for outcomes such as hospital readmission, mortality, and high-cost utilization relative to conventional statistical models. However, most of these studies focus on narrowly defined endpoints or single-disease cohorts. In contrast, this study addresses advanced chronicity, which inherently requires multidimensional assessment. The ability of the model to achieve high sensitivity is particularly relevant, as early identification of this cohort of patients remains a recognized gap in current care pathways, where detection is often delayed until late-stage deterioration or explicit palliative care referral.

From a clinical perspective, the implications of these findings are significant. First, the model demonstrates the feasibility of leveraging routinely available data to proactively identify patients who may benefit from anticipatory care planning, including palliative-oriented interventions. This approach could shift healthcare delivery from a reactive model to a more efficient and proactive framework, reducing time requirements relative to the current NECPAL-based assessment process and improving the detection of patients who might otherwise be overlooked. Second, the reliance on a limited number of interpretable variables facilitates their integration into clinical workflows. Third, the deliberate exclusion of variables directly linked to prior palliative care engagement ensures that the model is not merely reproducing existing clinical decisions, but instead identifies unrecognised cases, which is essential for real-world utility.

Nevertheless, several limitations must be acknowledged, and they materially affect the interpretation and applicability of the results. The most critical constraint is the small sample size (n = 163). Although feature selection and cross-validation were applied to mitigate overfitting, the risk of optimistic performance remains substantial. On the other hand, the near-balanced class distribution, while advantageous for model training, does not reflect real-world prevalence of advanced chronicity and may lead to miscalibration when applied in practice. Additional methodological limitations include the use of imputation strategies for missing data, which may underestimate variability and introduce bias. Finally, bias may also have been introduced through the exclusion of specific hospital services, although this was necessary to maintain conceptual consistency in the definition of advanced chronicity.

Future research should prioritize external validation in independent, multicentre cohorts to assess generalizability and robustness across different healthcare environments, and with real prevalence. Larger sample sized would enable the exploration of more complex modelling approaches as temporal models and DL architectures. The incorporation of longitudinal data could allow for dynamic risk prediction, capturing trajectories of decline rather than static snapshots. Finally, although the results reported here are promising, it will be essential to conduct a broader prospective clinical study to determine whether the implementation of such models translates into better patient outcomes, more appropriate resource allocation, and significant changes in clinical decision-making.

## 5. Conclusions

In this study, we present a high-performing ML-based approach for the earlier identification of patients with advanced chronic conditions. Among the evaluated algorithms, the bagging classifier demonstrated the most consistent overall performance, combining high sensitivity with strong discriminative capacity across all validation procedures. Notably, a reduced set of ten clinically meaningful variables, reflecting functional dependency, frailty, and healthcare utilization, was sufficient to achieve robust predictive performance, underscoring the relevance of these dimensions in characterizing advanced chronicity.

The model maintained high performance without incorporating variables directly linked to prior palliative care identification processes, thereby avoiding circular dependencies. This design choice enabled the detection of patients not previously identified through existing clinical pathways, reinforcing the potential of the model as a complementary screening tool rather than a confirmatory mechanism. Performance stability across CV further supports the internal robustness of the proposed approach, with consistent results observed across data partitions.

These findings demonstrate the feasibility of leveraging structured clinical data, complemented by relevant information extracted from clinical text notes, to support the early identification of MACA patients within a hospital setting. The observed performance, combined with the interpretability of the selected predictors, highlights the potential and clinical utility of the model as a decision-support tool for proactive care stratification.

As a next step, it is essential to evaluate the generalizability of the model in a prospective, real-world clinical study. This will allow assessment of its performance under true prevalence conditions, across different patient populations and clinical settings, and will provide critical evidence regarding its stability, transportability, and potential impact on clinical decision-making.

## Data Availability

All data produced in the present work are contained in the manuscript

## References

Barnett, K., Mercer, S. W., Norbury, M., Watt, G., Wyke, S., & Guthrie, B. (2012). Epidemiology of multimorbidity and implications for health care, research, and medical education: a cross-sectional study. Lancet (London, England), 380(9836), 37–43. 10.1016/S0140-6736(12)60240-2.

Bernabeu-Wittel, M., Ollero-Baturone, M., Moreno-Gaviño, L., Barón-Franco, B., Fuertes, A., Murcia-Zaragoza, J., Ramos-Cantos, C., Alemán, A., & Fernández-Moyano, A. (2011). Development of a new predictive model for polypathological patients. The PROFUND index. European journal of internal medicine, 22(3), 311–317. 10.1016/j.ejim.2010.11.012.

Billings, J., Blunt, I., Steventon, A., Georghiou, T., Lewis, G., & Bardsley, M. (2012). Development of a predictive model to identify inpatients at risk of re-admission within 30 days of discharge (PARR-30). BMJ open, 2(4), e001667. 10.1136/bmjopen-2012-001667.

Boyd, C. M., Darer, J., Boult, C., Fried, L. P., Boult, L., & Wu, A. W. (2005). Clinical practice guidelines and quality of care for older patients with multiple comorbid diseases: implications for pay for performance. JAMA, 294(6), 716–724. 10.1001/jama.294.6.716.

Charlson, M. E., Pompei, P., Ales, K. L., & MacKenzie, C. R. (1987). A new method of classifying prognostic comorbidity in longitudinal studies: development and validation. Journal of chronic diseases, 40(5), 373–383. 10.1016/0021-9681(87)90171-8.

Futoma, J., Morris, J., & Lucas, J. (2015). A comparison of models for predicting early hospital readmissions. Journal of biomedical informatics, 56, 229–238. 10.1016/j.jbi.2015.05.016.

Goldstein, B. A., Navar, A. M., & Carter, R. E. (2017). Moving beyond regression techniques in cardiovascular risk prediction: applying machine learning to address analytic challenges. European heart journal, 38(23), 1805–1814. 10.1093/eurheartj/ehw302.

Gómez-Batiste, X., Martínez-Muñoz, M., Blay, C., Amblàs, J., Vila, L., Costa, X., Villanueva, A., Espaulella, J., Espinosa, J., Figuerola, M., & Constante, C. (2013). Identifying patients with chronic conditions in need of palliative care in the general population: development of the NECPAL tool and preliminary prevalence rates in Catalonia. BMJ supportive & palliative care, 3(3), 300–308. 10.1136/bmjspcare-2012-000211.

Hippisley-Cox, J., & Coupland, C. (2017). Development and validation of QMortality risk prediction algorithm to estimate short term risk of death and assess frailty: cohort study. BMJ (Clinical research ed.), 358, j4208. 10.1136/bmj.j4208.

Jiménez, I., Pajarón, M., Moreno-Ariño, M., Campmol, M. & Monte, R. (2021). Prevalence and Mortality of Patients with Palliative Needs in an Acute Respiratory Setting. Archivos de Bronconeumología (English Edition). 57. 10.1016/j.arbr.2021.09.010.

Kansagara, D., Englander, H., Salanitro, A., Kagen, D., Theobald, C., Freeman, M., & Kripalani, S. (2011). Risk prediction models for hospital readmission: a systematic review. JAMA, 306(15), 1688–1698. 10.1001/jama.2011.1515.

Kelly, C. J., Karthikesalingam, A., Suleyman, M., Corrado, G., & King, D. (2019). Key challenges for delivering clinical impact with artificial intelligence. BMC medicine, 17(1), 195. 10.1186/s12916-019-1426-2.

Krumholz H. M. (2014). Big data and new knowledge in medicine: the thinking, training, and tools needed for a learning health system. Health affairs (Project Hope), 33(7), 1163–1170. 10.1377/hlthaff.2014.0053.

Lunney, J. R., Lynn, J., Foley, D. J., Lipson, S., & Guralnik, J. M. (2003). Patterns of functional decline at the end of life. JAMA, 289(18), 2387–2392. 10.1001/jama.289.18.2387.

Lynn, J. & Adamson, DM. (2003). Living Well at the End of Life: Adapting Health Care to Serious Chronic Illness in Old Age. Santa Monica, CA: RAND Corporation. https://www.rand.org/pubs/white_papers/WP137.html.

Miotto, R., Li, L., Kidd, B. et al. (2016). Deep Patient: An Unsupervised Representation to Predict the Future of Patients from the Electronic Health Records. Sci Rep 6, 26094. 10.1038/srep26094

Murray, S. A., Kendall, M., Boyd, K., & Sheikh, A. (2005). Illness trajectories and palliative care. BMJ (Clinical research ed.), 330(7498), 1007–1011. 10.1136/bmj.330.7498.1007.

Obermeyer, Z., & Emanuel, E. J. (2016). Predicting the Future - Big Data, Machine Learning, and Clinical Medicine. The New England journal of medicine, 375(13), 1216–1219. 10.1056/NEJMp1606181.

Obermeyer, Z., Powers, B., Vogeli, C. & Mullainathan, S. (2019). Dissecting racial bias in an algorithm used to manage the health of populations. Science 366, 447–453 (2019). DOI: 10.1126/science.aax234

Rajkomar, A., Oren, E., Chen, K. et al. Scalable and accurate deep learning with electronic health records. npj Digital Med 1, 18 (2018). 10.1038/s41746-018-0029-1.

Shickel, B., Tighe, P. J., Bihorac, A., & Rashidi, P. (2018). Deep EHR: A Survey of Recent Advances in Deep Learning Techniques for Electronic Health Record (EHR) Analysis. IEEE journal of biomedical and health informatics, 22(5), 1589–1604. 10.1109/JBHI.2017.2767063.

Smith, S. M., Wallace, E., O’Dowd, T., & Fortin, M. (2016). Interventions for improving outcomes in patients with multimorbidity in primary care and community settings. The Cochrane database of systematic reviews, 3(3), CD006560. 10.1002/14651858.CD006560.pub3.

Temel, J. S., Greer, J. A., Muzikansky, A., Gallagher, E. R., Admane, S., Jackson, V. A., Dahlin, C. M., Blinderman, C. D., Jacobsen, J., Pirl, W. F., Billings, J. A., & Lynch, T. J. (2010). Early palliative care for patients with metastatic non-small-cell lung cancer. The New England journal of medicine, 363(8), 733–742. 10.1056/NEJMoa1000678.

Violan, C., Foguet-Boreu, Q., Flores-Mateo, G., Salisbury, C., Blom, J., Freitag, M., Glynn, L., Muth, C., & Valderas, J. M. (2014). Prevalence, determinants and patterns of multimorbidity in primary care: a systematic review of observational studies. PloS one, 9(7), e102149. 10.1371/journal.pone.0102149.

Wynants, L., Van Calster, B., Collins, G. S., Riley, R. D., Heinze, G., Schuit, E., Bonten, M. M. J., Dahly, D. L., Damen, J. A. A., Debray, T. P. A., de Jong, V. M. T., De Vos, M., Dhiman, P., Haller, M. C., Harhay, M. O., Henckaerts, L., Heus, P., Kammer, M., Kreuzberger, N., Lohmann, A., … van Smeden, M. (2020). Prediction models for diagnosis and prognosis of covid-19: systematic review and critical appraisal. BMJ (Clinical research ed.), 369, m1328. 10.1136/bmj.m1328.

